# A cohort study on the duration of Plasmodium falciparum infections during the dry season in The Gambia

**DOI:** 10.1101/2021.11.12.21266275

**Authors:** Katharine A Collins, Sukai Ceesay, Sainabou Drammeh, Fatou K Jaiteh, Marc-Antoine Guery, Kjerstin Lanke, Lynn Grignard, Will Stone, David J Conway, Umberto D’Alessandro, Teun Bousema, Antoine Claessens

## Abstract

**Background:** In areas where *Plasmodium falciparum* malaria is highly seasonal, a dry season reservoir of blood-stage infection is essential for initiating transmission during the following wet season, bridging transmission seasons several months apart. Understanding infections during the dry season could thus inform approaches for malaria control.

**Methods:** In The Gambia, a cohort of 42 individuals with qPCR positive *P. falciparum* infections at the end of the transmission season (December) were followed monthly until the end of the dry season (May) to evaluate the duration of detectable infections. The influence of human host (age, sex, haemoglobin concentration and genotype, and *P. falciparum*-specific antibodies), and parasitological (parasite density, gametocyte density and genotypic multiplicity of infection) factors was investigated.

**Results:** A large proportion of individuals infected at the end of the wet season had detectable infections until the end of the dry season (40.0%; 16/40), with the majority of these infections also harbouring gametocytes (81.3%; 13/16). 22 infections were classified as persistent (detectable for at least 3 months), 17 were classified as short-lived (undetectable within 2 months), and 3 were treated (due to symptoms). At the start of the dry season, the majority of persistent infections (82%; 18/22) had parasite densities >10 p/µL compared to only 5.9% (1/17) of short-lived infections. Persistent infections (59%; 13/22) were also more likely to be multi-clonal than short-lived infections (5.9%; 1/17), they were most common in 5 to 15 year old children (63%; 12/19), and were associated with individuals having higher levels of *P. falciparum*-specific antibodies (p = 0.058).

**Conclusions:** Asymptomatic persistent dry season infections in The Gambia were multiclonal with higher parasite densities at the beginning of the dry season, mostly occurring in school age children and adults with higher *P. falciparum-*specific antibodies. Screening and treating asymptomatic, malaria-infected individuals during the dry season may reduce the human reservoir of malaria responsible initiating transmission in the wet-season.

## Introduction

In many endemic areas, malaria transmission is seasonal, with most clinical cases occurring during the wet season. *Plasmodium falciparum* infections acquired during this period can persist in asymptomatic individuals throughout the dry season, and if they produce gametocytes, can potentially contribute to transmission at the start of the next wet season, bridging transmission seasons several months apart [1-3]. These clinically silent infections can persist for many months or even years without treatment [2], and pose a significant hurdle for malaria elimination efforts [4]. Understanding how long infections persist in the dry season in The Gambia and identifying who carries them could inform targeted approaches to reduce the dry season reservoir of malaria and reduce re-introduction of malaria in the subsequent wet season.

Few studies have described in detail the asymptomatic dry season reservoir of malaria and the related parasite and host factors. Duration of *P. falciparum* infection varies by setting, with asymptomatic, genetically complex infections being more common in high transmission areas [5], but even in low transmission settings infections are known to persist for months or even years [6, 7]. The host specific response to malaria infection may play an important role by allowing the establishment of chronic infections. Protection against severe malaria is acquired very quickly, often after a single infection [8, 9], but the ability to control parasitemia at a sub-clinical level develops more slowly with age, following repeated exposure [10]. In Mali, individuals infected during the dry season had higher *P. falciparum-*specific antibodies and memory B cells than those who were uninfected [11, 12], and older children were more likely to be infected at the end of the dry season than younger children [13]. Both findings support the hypothesis that a certain degree of exposure and immunity (acquired with increasing age) is required to be asymptomatically infected. Interestingly, in Uganda, older children carried infections for a longer time than children under 5 and adults [14], suggesting host immunity may peak or plateau at a certain age, or there may be different mechanisms involved as immunity develops. The type of immune response associated with persistent asymptomatic infection may differ from that able to clear an infection, and this may also vary by age and exposure [15]. Moreover, it is unclear when parasite specific antibody responses contribute to protection [16, 17] or are simply markers of previous exposure [18]. In a recent study, responses to a panel of *P. falciparum* antigens indicated a higher level of immunity that allowed individuals to control blood-stage infections following controlled exposure to *P. falciparum* sporozoites [19]. Other host factors may also play an important role in the establishment of chronic asymptomatic infections. In Uganda, infections persisted longer in males than females, suggesting sex-specific differences in the host response may influence the duration of parasite carriage [14]. Host factors such as haemoglobinopathies [20] and anaemia [21] are known to be associated with the risk and severity of malaria, and may also be important factors in the duration of asymptomatic carriage [22].

Besides host factors related to chronic infections, parasites themselves may either be pre-programed or adapted to persist throughout the dry, non-transmission season. Being able to establish a clinically silent infection that does not activate the immune system may result in a slower clearance rate, a plausible evolutionary strategy for surviving the dry season and ensuring onward transmission. In Mali, where malaria transmission is highly seasonal, parasite populations collected during the wet and dry seasons were similar. Nevertheless, parasites sampled in the dry season appeared to adapt their phenotype, resulting in a longer time in peripheral circulation with increased splenic clearance, potentially a strategy to maintain a clinically and immunologically silent low parasitemia [11]. In this study, we identified individuals infected at the end of the transmission season in The Gambia, determined the duration of these infections during the dry season, and explored the related host and parasite factors.

## Methods

### Study design

Climate in The Gambia is characterised by a short rainy season and a long dry season. Malaria transmission follows the same pattern, starting in July or August and lasting until December, with little or no clinical cases during the dry season. In December 2016, all residents of four villages (Madina Samako, Sendebu, Njayel, Karanbada) located within 5Km of each other in the Upper River Region, eastern Gambia, had a finger prick blood sample collected and analysed by VarATS quantitative PCR (qPCR) for *P. falciparum* infection. qPCR-positive individuals were invited for monthly samplings (venous blood draw of 5 to 8mL), with qPCR and qRT-PCR performed from the end of December until the end of the dry season (May), or until they either, (i) they became symptomatic and received treatment immediately, (ii) cleared their infection (negative by qRT-PCR and/or VarATS qPCR two months in a row), or (iii) withdrew from the study (Figure 1a). Subjects with an axillary temperature greater than 37.5°C were screened with a rapid diagnostic test (CareStart) and positives were treated with artemether-lumefantrine, the first line treatment in The Gambia. Written informed consent was obtained from all participants; parents or legal guardians provided a written informed consent for minors (<18 years old). The study was approved by the Gambia Government/Medical Research Council Joint Ethics Committee (SCC 1318, L2014.67, SCC 1472).

**Figure 1:**
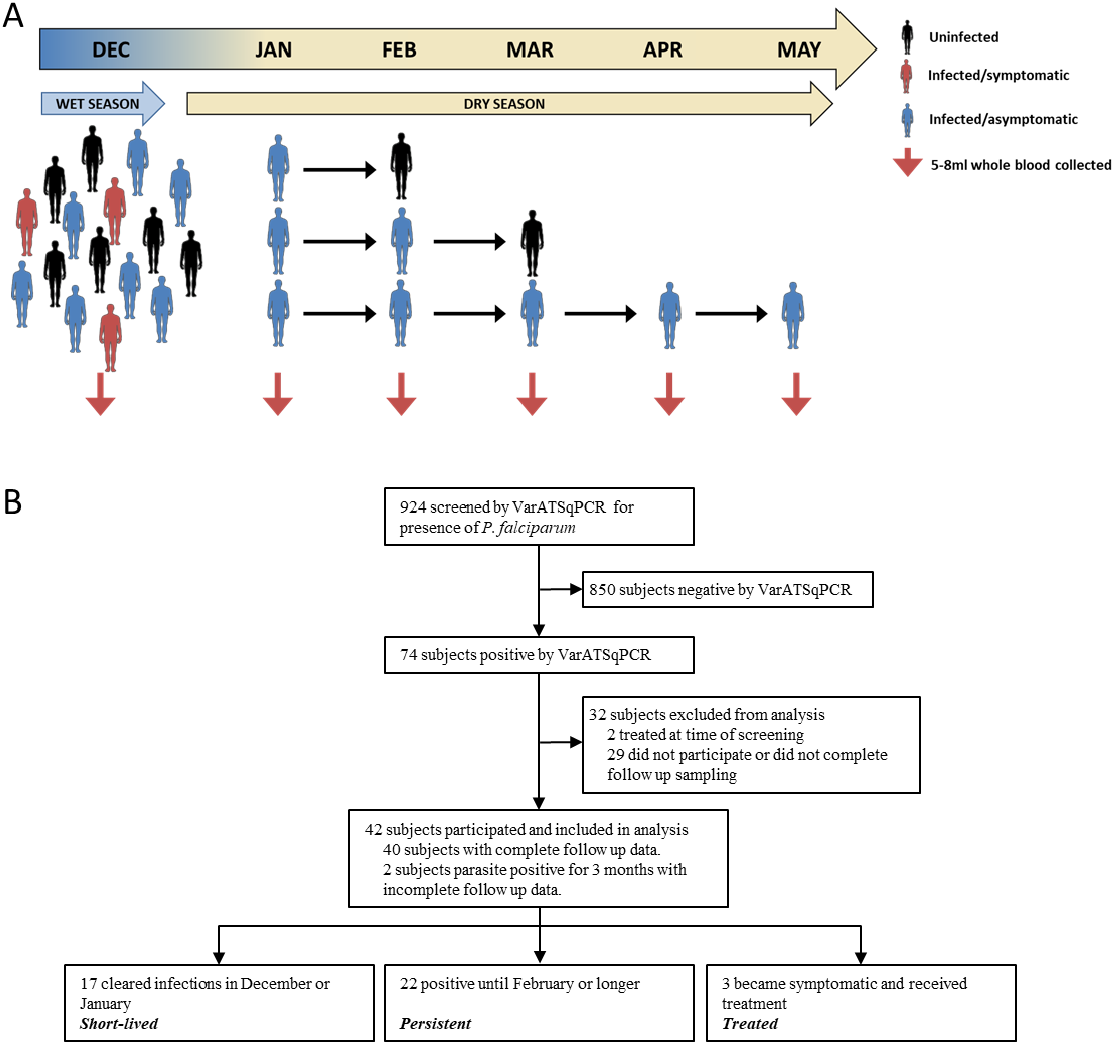
Study design and flow. (A) Illustration of study design (B) Study flow diagram. Subjects were screened in December 2016 and asymptomatic parasite positive individuals (by VarATS qPCR) were selected at the end of the wet season (December 2016) and were recruited into a longitudinal study with monthly samples to determine duration of infection from end of December until the end of the dry season (May) or until they (i) cleared their infection (negative by PCR), or (ii) became symptomatic and received treatment.

### Molecular detection of parasites

During the study, presence of *P. falciparum* was assessed using VarATS qPCR as described in [23] from fingerprick (start December screening) or venous blood samples (monthly end December to May). The parasite stages were determined by qRT-PCR assays specific for ring stage parasites (*sbp-1)*, male gametocytes (*PfMGET*), or female gametocytes (*CCp4*) [24]. Samples were genotyped using *P. falciparum* merozoite surface protein 2 *(msp2)* [25]. Briefly, nucleic acid was extracted from 100 µl whole blood and 5 µl was used in a nested PCR specific for *msp2* 3D7 and FC27 allelic families followed by capillary electrophoresis to determine allelic size polymorphisms. Secondary PCR products were run on a 2% w/v agarose gel to check for bands within the correct size range (193 to 506 bp). Secondary PCR products were diluted and 2.5 µl per sample processed on an ABI 7500 fast (Applied Biosystem, UK) and resulting chromatograms analyzed using PeakScanner® software version 2 (Applied Biosystems). To distinguish between true peaks and background signal or stutter peaks, a cutoff of 200 relative fluorescent units (RFUs) was applied and fragment sizes must differ by more than 3 bp.

### Haemoglobin quantification and genotyping

Haemoglobin density was measured in December using a haemoglobin analyser (HemoCue; AB Leo Diagnostics, Helsingborg, Sweden). Human haemoglobin S (HBs) and C (HBc) were genotyped using previously published methods [26].

### Luminex

IgG antibodies against 17 *P. falciparum* antigens were quantified at baseline for each participant using a Luminex MAGPIX© suspension bead array [27]: Circumsporozoite protein (CSP), Erythrocyte binding antigen (EBA140, EBA175 and EBA181); Glutamate rich protein 2 (GLURP-R2); Merozoite surface protein 1-19 (MSP1-19), Merozoite surface protein 2 (MSP2-ch150/9 [3D7 family allele] and MSP2-DD2 [FC27 family allele]); Apical membrane antigen 1 (AMA1), Reticulocyte binding protein homologue (RH2.2, RH4.2, RH5.1), Schizont egress antigen-1 (SEA1), Heat shock protein 40 (HSP40), Skeleton-binding protein 1 (SBP1), and Early transcribed membrane proteins 4 (ETRAMP4 Ag2) and 5 (ETRAMP5 Ag1). Full details of all antigens are in supplementary table 1. These antigens are wither expressed during the asexual blood-stage (Etramp, PfSEA, HSP40, SBP1), involved in merozoite invasion (Rh, AMA1, EBA, MSP, GLURP) or localised on the surface of sporozoites (CSP). Serological analysis was conducted on serum samples collected in December, assayed at a dilution of 1:200. Secondary antibody was an R-phycoerythrin conjugated goat anti-human IgG (Jackson Immuno Research, PA, USA; 109-116-098) diluted 1:200. Data are presented as median fluorescence intensity (MFI) (median of individual bead fluorescence values for a given region/antigen specificity) adjusted for inter-plate variation in background reactivity as previously described [27]

### Statistical analysis

Infections were classified as ‘short-lived’ if they were cleared within 2 months, ‘persistent’ if they remained detectable for 3-6 months and ‘treated’ if they became symptomatic and were treated. Data were analysed with Graphpad Prism version 9. A normality test (D’Agostino-Pearson) was used to determine if data were normally distributed. Kruskal-Wallis with Dunns multiple comparison test was used to compare 3 or more groups of non-parametric data. Spearmans correlation was used to analyse associations with non-parametric data and Pearsons correlation was used for parametric data. Significance was indicated when the value of p < 0.05 (*p < 0.05, **p < 0.01, ***p < 0.001).

## Results

Malaria prevalence at the end of the transmission season (beginning of December 2016) was 8.0% (74/924), with children under 5 having lower prevalence (2.5%; 5/199) than children 5-15 years old (7.9%; 27/341) and individuals >15 years old (10.9%; 42/384). Complete follow-up data were obtained from 40 individuals, 2 did not complete follow up until negative by PCR, but were parasite positive for at least 3 months so were included in the categorised analysis only (Figure 1a and b). 41% (n=17) of the subjects had *short lived* infections (undetectable by January), 52% (n=22) had *persistent* infections (detectable by PCR for 3-6 months), and 3 infected subjects developed symptoms and were *treated* (7%, all children <15 years of age) (Figure 1b). 40% (16/40) of individuals infected in early December remained PCR positive until the end of the dry season, with most of these individuals (81.3%; 13/16) also carrying gametocytes at the end of the dry season (Figure 2a).

**Figure 2.**
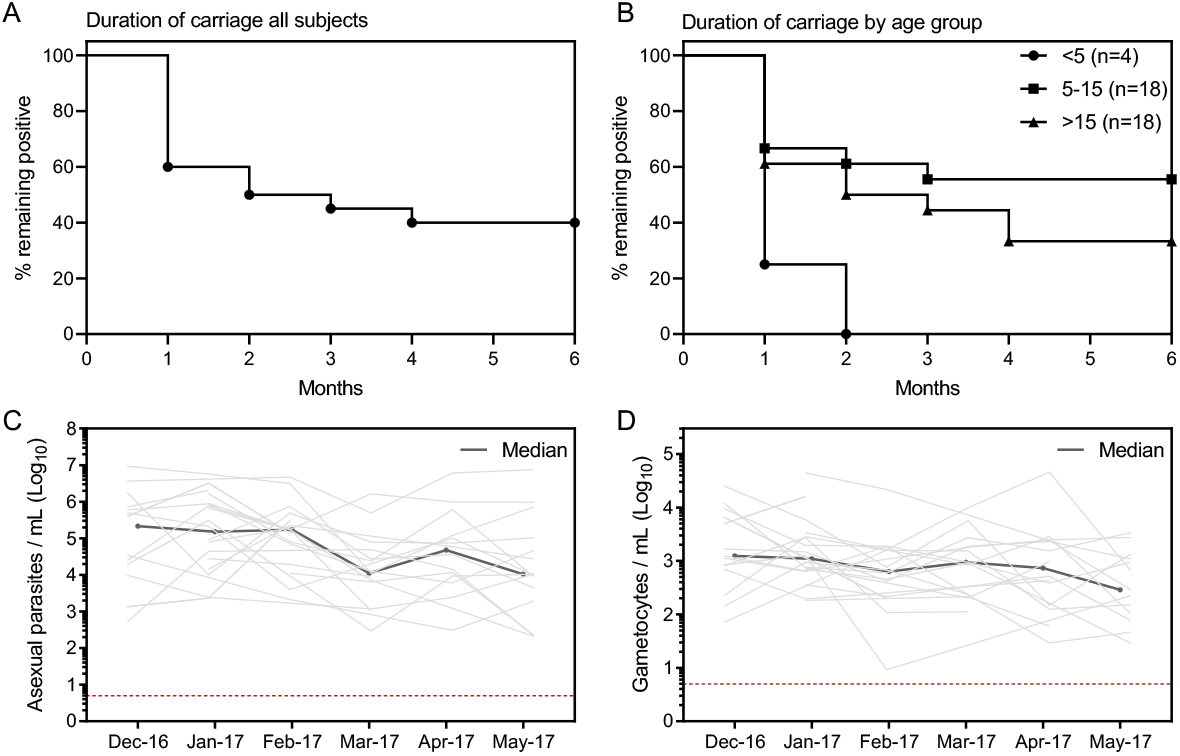
Duration of detectable parasite carriage in the blood during the dry season. Subjects (n=42) who were parasite positive by VarATS qPCR in fingerprick blood samples in December were followed monthly until PCR negative or treated. Two subjects who did not complete follow up were parasite positive for at least 3 months (and thus classified as persistent infections) are excluded from the Kaplan Meier graphs). (A) Number of subjects parasite positive each month (n=40). (B) Number of subjects positive each month by age category. (C) Asexual parasite densities over time in the persistent infections group, measured in venous blood samples by qRT-PCR for ring stage parasite (*SBP-1*). (D) Total gametocyte densities (males + females) over time in the persistent infections group, measured in venous blood samples by qRT-PCR for male (*PfMGET*) and female (*CCp4*) gametocytes. Light grey lines show individual responses and the dark grey line shows the median response.

No association between the sex of the individual and the duration of infection was found (Table S2). Mean haemoglobin concentration at the start of the dry season was 11.31 g/dL (n=37). HbAA was the most frequent haemoglobin type (75.7%; 28/37), followed by HbAS (21.6%, 8/37), and HbSS (2.7%, 1/37). There was no association between haemoglobin concentration (short-lived= 11.35, persistent = 11.18, treated = 11.30) or type and duration of carriage (Table S2). Persistent infections were slightly more common in the 5-15 year olds (63%; 12/19) than in the other age groups (<5years - 0.0% [0/4], >15 years - 53% [10/19]) (Figure 2b). In persistent infections, asexual parasitemia and gametocytemia declined slowly over the season, with the median asexual parasitemia declining from 214,417 to 10,446 parasites/mL (Figure 2c) and gametocytemia declining from 1,237 to 289 gametocytes/mL (Figure 2d). The ratio of male:female gametocytes was female biased (0.20; CI 95%:0.16-0.23), similar to sex ratios observed in transmissible infections [4] and did not change over time (Figure S2).

At the start of the dry season, parasite density in persistent infections was significantly higher than in short-lived infections (p< 0.0001 Kruskal-Wallis with Dunns multiple comparison test) (Figure 3a), and duration of infection in months was significantly related to starting parasite density (Spearmans r = 0.86, p< 0.0001) (Figure 3b). Most persistent infections (82%; 18/22) and only 1 out of 17 short-lived infections had initial parasite densities above the theoretical limit of expert microscopy detection (10p/uL) [28] (Figure 3a). In the end of December samples (first monthly visit after screening), most short-lived infections (76%; 13/17) had only gametocytes, and 53% (9/17) had neither asexual parasites nor gametocytes detected, indicating these individuals had already cleared their infections in the first month. Whereas at the same time point, almost all persistent infections had both asexual parasites (20/22) and gametocytes (20/22) at densities that could be quantified by the qRT-PCR assays.

**Figure 3.**
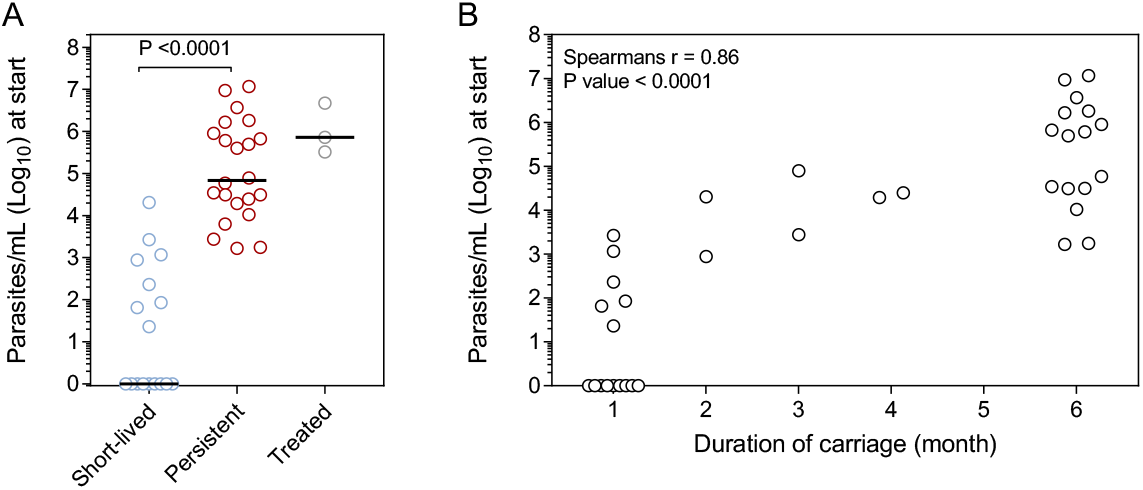
Level of parasitemia at the beginning of the dry season and subsequent duration of detectable parasitemia. Total parasite density (sum of asexual parasites and male and female gametocytes) for each subject (n=42) by qRT-PCR at the start of the dry season. (A) Total parasite density for each individual, by group. Groups compared by Kruskal-Wallis with Dunns multiple comparison test. (B) Spearmans correlation between parasite density and duration of infection for the persistent and short-lived infection groups.

Complexity of infection (the number of genetically distinct genotypes within a blood sample) was measured throughout the dry season using *msp2* genotyping (Figure 4a). In some samples, *msp2* genotype could not be determined, likely due to parasite densities being below the assay limit of detection. At the start of the dry season, most infections had less than 2 genotypes (80.5%, 33/41) and the median per subject was 1 (range 1-8). Complexity of infection was significantly higher in the persistent than in the short-lived infections (p = 0.015, Kruskal-Wallis with Dunns multiple comparison test). Higher complexity of infection (>2 genotypes) was only observed in persistent infections; all short-lived infections were monoclonal, except for one with 2 genotypes (Figure 4b). The number of genotypes at the start of the dry season also correlated with parasite density (Figure 4c). In every persistent infection, there was at least one *msp2* genotype detected repeatedly throughout the dry season, suggesting chronic infection with the same *P. falciparum* strain as opposed to new infections (Figure S3).

**Figure 4.**
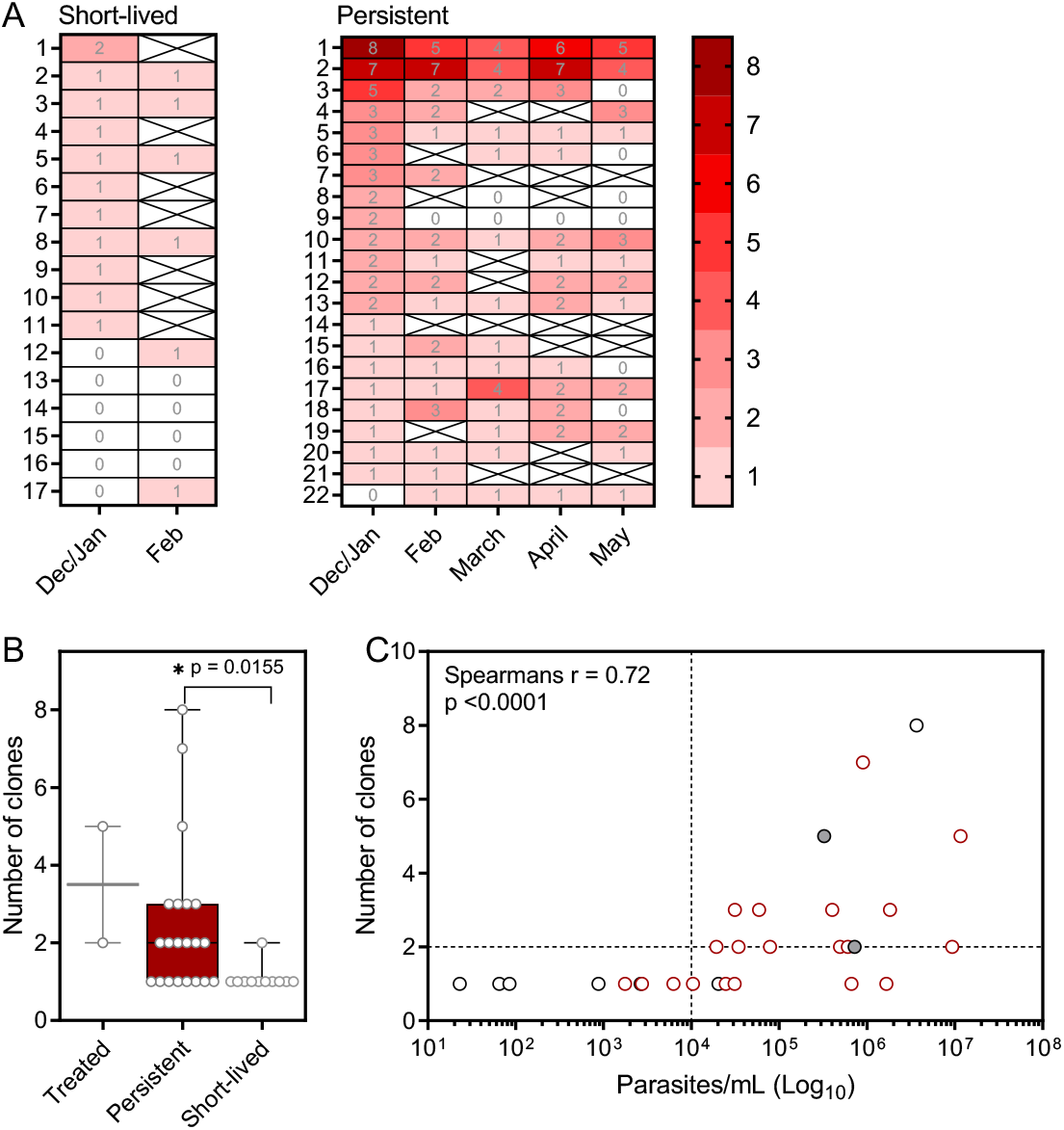
Genotypic complexity of parasite infection and persistent detectability through the dry season. The number of clones present in each individual was determined by *msp2* genotyping. (A) Number of clones for each individual over time, displayed by group. The black cross indicates where a sample was missing and a ‘0’ indicates the number of genotypes could not be determined. (B) Number of clones at the start of the dry season in the December or January sample, by group. For 7 subjects there is no genotyping data available from December or January. Groups compared by Kruskal-Wallis with Dunns multiple comparison test. (C) Spearman’s correlation between number of clones and total parasite density at the start of the dry season (sum or asexual parasites and gametocytes determined by qRT-PCR). Persistent infections (red circles), short-lived infections (black circles), treated (filled circles). Horizontal dashed line indicated the theoretical limit of expert microscopy detection

To assess the role of host immunity in the persistence of *P. falciparum* infection, antibodies were measured at the start of the dry season using a multiplex bead-based assay with a panel of 17 *P. falciparum* antigens. There was a trend for higher *P*.*f*.-specific antibody responses against 13 out of the 17 antigens in subjects who developed persistent infections compared to those with short-lived infections (Figure 5a and b). The three Rh family antigens (Rh2, Rh4.2, Rh5.1) were among the four most discriminatory antigens, with higher antibody levels associated with persistent infections, although only Rh5.1 reached statistical significance (p = 0.03). Of the other individual antigens, only PfSEA and GLURP (p = 0.01, p = 0.04, respectively) were significantly associated with subsequent duration of infection (persistent versus short-lived). Individuals with persistent infections tended to respond to more antigens (above background response) than those with short-lived infections, p = 0.058 (Figure 5c). The magnitude of total *P*.*f*.-specific antibody response to all antigens was significantly correlated with age, Pearsons r = 0.43 p = 0.017. (Figure 5d) and there was also a trend for increased breadth of response with age (Figure 5e).

**Figure 5.**
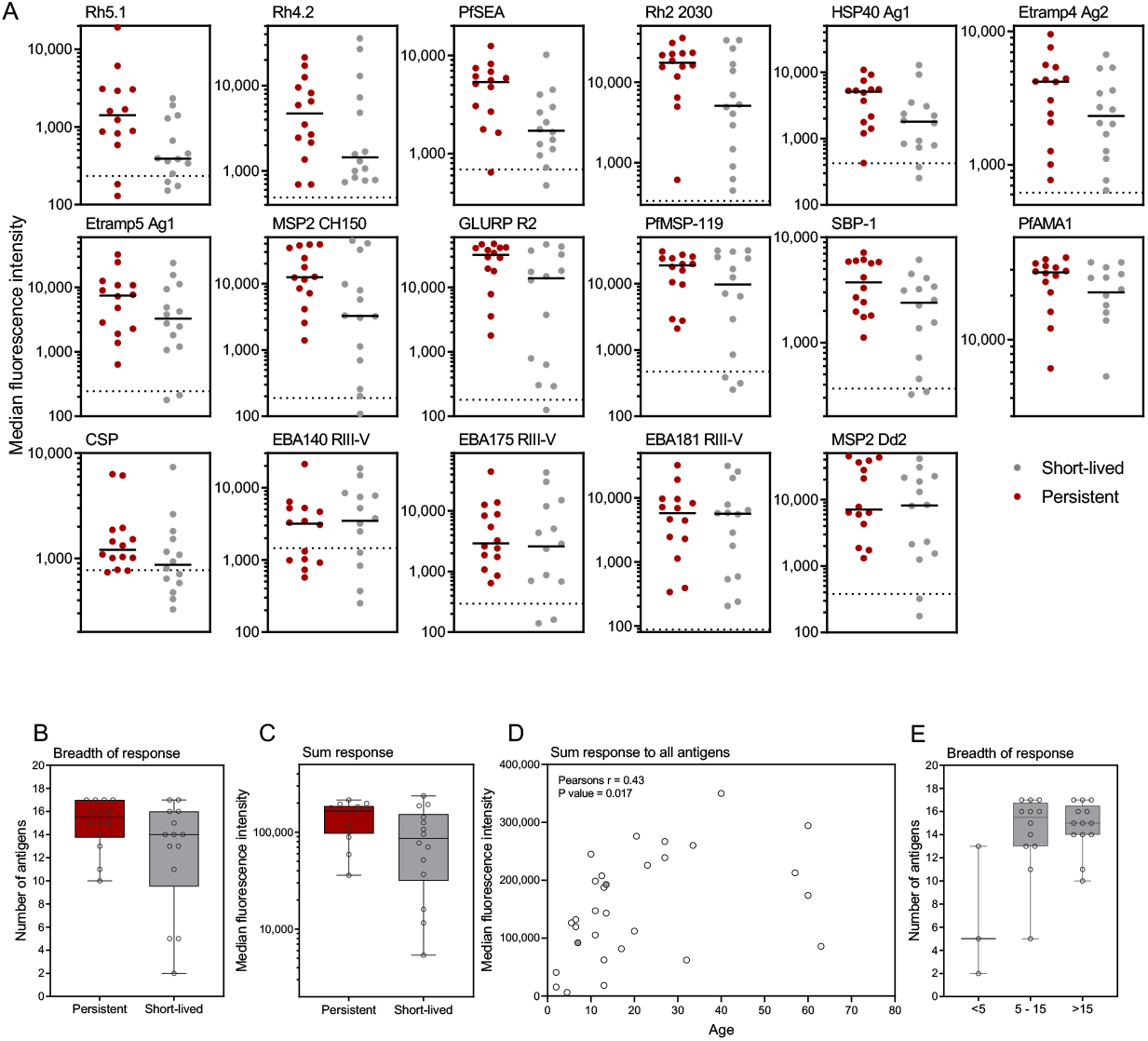
Antibody levels to a panel of *P. falciparum* antigens in relation to duration of detectable infection in the dry season. Antibody responses were measured to a panel of 17 *P. falciparum* antigens using a multiplex bead-based assay. (A) Panels show the individual subject responses to each antigen, by short-lived or persistent infection group, ordered by decreasing differences between the two groups. Dashed line indicates average background response. (B) Sum of the response all 13 upregulated antigens by e group. (C) Number of antigens each individual responded to above background by group. (D) Pearsons correlation between age and total antibody response to all 17 antigens (treated - grey filled circles). (E) Number of antigens each individual responded to above background by age group. Box plots show the median with the whiskers indicating the minimum and maximum

## Discussion

In this study, 40% of individuals infected at the end of the wet season remained asymptomatically infected until the end of the dry season, demonstrating the considerable silent reservoir of malaria that exists to re-start the next transmission season in The Gambia. Individuals with persistent infections had higher starting parasite densities, often with multiclonal infections, and higher *P*.*f*.-specific antibody responses, suggesting that prior malaria exposure is important to maintain the human reservoir of malaria. Targeting these infections may significantly reduce such a reservoir and thus the capacity of starting the next transmission season.

In areas of seasonal malaria transmission, understanding who maintains the parasite reservoir during the dry season may inform control interventions to reduce this reservoir and thereby the resurgence of malaria in the subsequent transmission season [1]. In our study setting, only 8% of village residents had detectable parasites at the end of the transmission season and towards the end of the dry season the prevalence fell to 3%. In our modestly sized study population, we aimed to understand factors that contribute to duration of infection. We observed no clear relationship between subject age and duration of parasite carriage, although all persistent infections were found in those above 5 years of age. In general, the number of children under 5 infected at the end of the wet season was low compared to the other age groups, possibly reflecting the levels of protective immunity, with younger children being more likely to develop symptoms when infected [7], or the impact of seasonal malaria chemoprevention [29].

Responses to the panel of 17 *P. falciparum* antigens tested were generally low [18]. Despite being currently infected, not all subjects had a response to all antigens above background. This highlights the generally low level of malaria exposure in the area (when compared to high endemic settings). Individuals with persistent infections tended to have higher and broader *P*.*f*.*-*specific antibody responses than those with short-lived infections, a finding consistent with previous studies. In Mali, a certain level of immunity was required for chronic asymptomatic parasite carriage when compared to uninfected individuals [11, 12]. Similarly, in another study in Mali, asymptomatic infected children had higher levels of *P*.*f*.-specific antibodies than children with clinical malaria [13]. Other studies have shown that age (and thus cumulative exposure) is associated with longer duration of asymptomatic infection in Malawi [30], and this relationship was seen particularly in children of different ages in Uganda and Ghana [14, 31]. In our study, we also observed a relationship between age and increasing magnitude and breadth of the immune responses, but this correlation was relatively weak, with some older individuals having poorer antibody responses. This supports earlier reports of marked heterogeneity in malaria exposure in The Gambia [3, 32]. In the context of our current findings, we hypothesize that this heterogeneity results in some individuals being more likely to experience repeated infections, resulting in higher levels of immunity and thus improved ability to sustain chronic asymptomatic parasite carriage. Due the heterogeneous nature of exposure, these individuals are more accurately characterised by responses to a panel of *P. falciparum* antigens than by age.

Each persistent infection had at least one unique genotype repeatedly observed at several time points, indicating infection persistence. High complexity of infection (multiple genotypes) at the beginning of the dry season was associated with longer duration of asymptomatic carriage, consistent with data from other studies [33, 34]. This could either be due to multiple clones being received from a single mosquito bite [35], or by the accumulation of clones over time in those who harbour infections for a prolonged period. Complexity of infection and parasite density were also strongly correlated in this study. This could be partly explained by the difficulty of detecting all parasite clones when density is low and falling below the detection limit of the assay [36]. Higher parasite density, multi-clonal infections were more likely to persist throughout the dry season. This could possible be because a higher parasite biomass in December would take longer to clear. However, this was not clearly supported by our data since parasite densities in persistent infections did not decrease dramatically during the dry season. Alternatively, parasites in persistent infections may be (epi)genetically programmed to survive longer, and/or genetically different from those in short-lived infections, potentially with a less virulent phenotype or slower parasite multiplication rate. This hypothesis is consistent with a recent study from Mali, where similar parasite populations were present in the wet and dry seasons. However, parasites in the dry season were transcriptionally different from those present in the wet season and they were able to adapt their phenotype to ensure longer circulation and less sequestration during each cycle, resulting in increased splenic clearance and lower but persistent parasite densities [11].

Our study was limited by the small sample size, largely governed by the decreasing malaria prevalence in the area [3]. However, we were able to demonstrate that the dry season reservoir of malaria is largely comprised of school aged children and adults who have likely experienced repeated infections. Importantly, persistent infections could potentially be identified at the start of the dry season, given that they were associated with higher initial parasite densities. Our findings confirm ongoing gametocyte production in chronic infections. Future studies may address the infectivity of these infections to mosquitoes and, more importantly for malaria control, determine whether clearing infections that are present at the start of the dry season results in a meaningful reduction in malaria burden in the next transmission season.

## Data Availability

All data produced in the present study are available upon request to the authors

## Supplementary Figures

**Figure S1.**
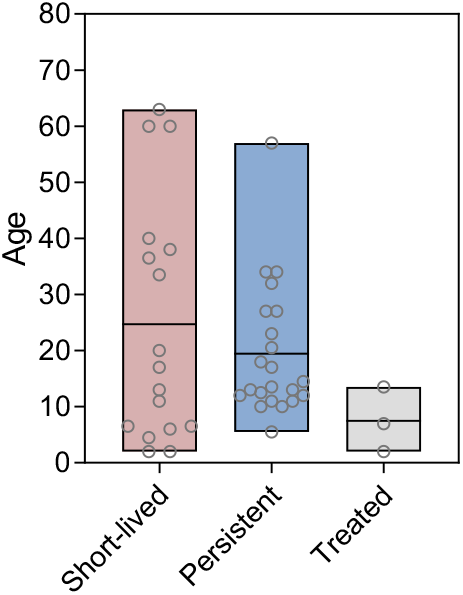
Age distribution and duration of carriage.

**Figure S2.**
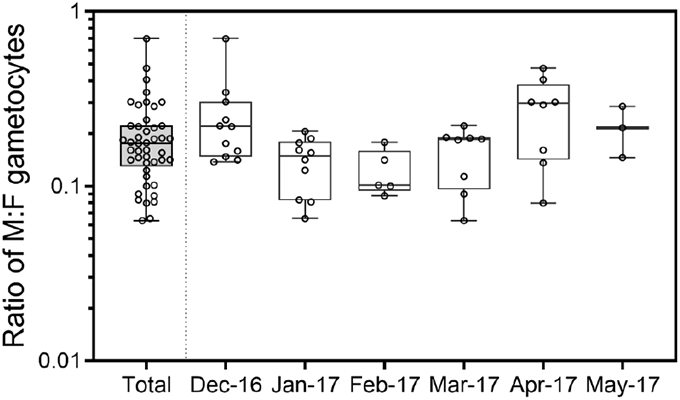
Sex ratio of gametocytes.

**Figure S3.**
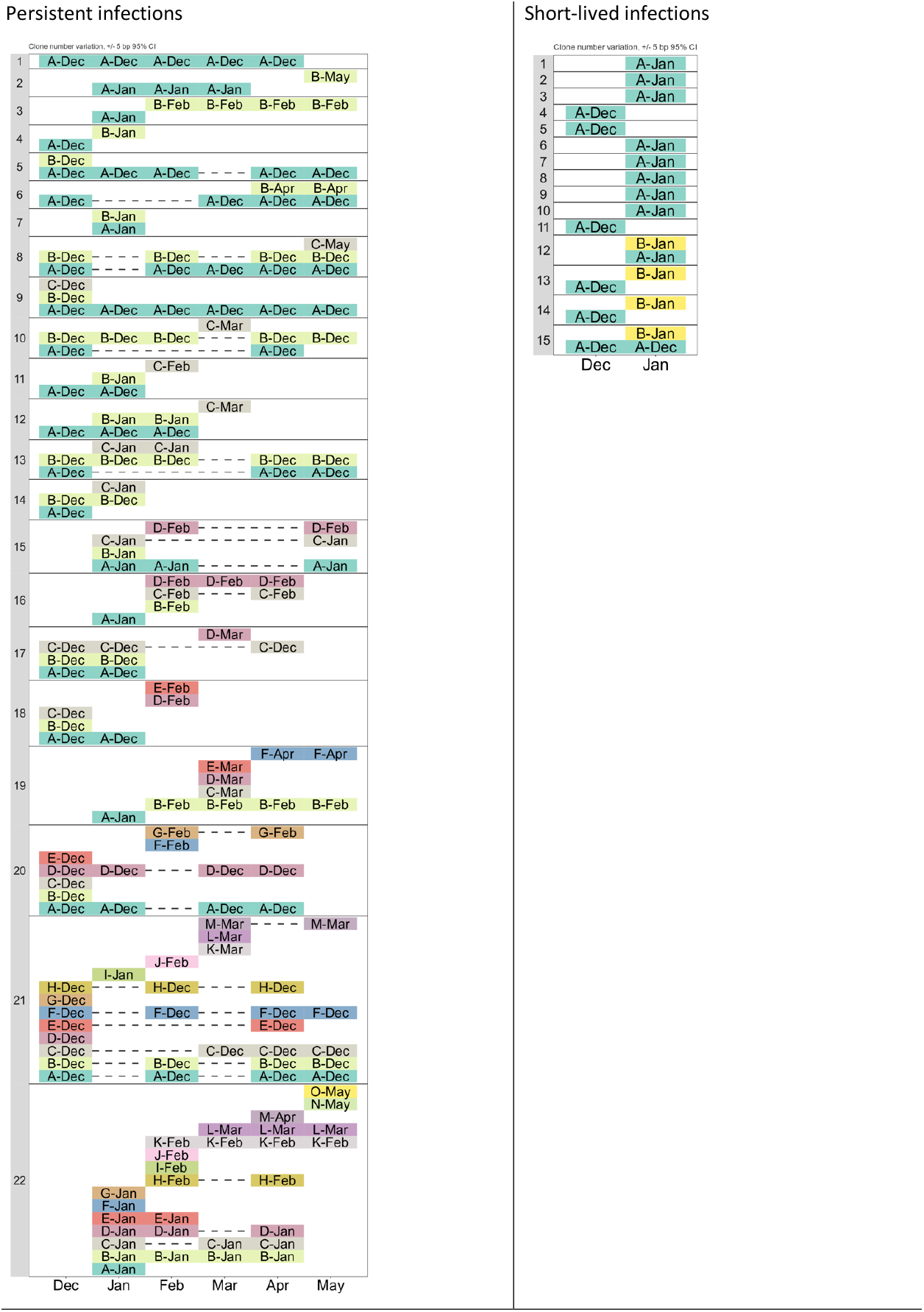
Number and types of *MSP2* genotypes per sample. In each volunteer (left-hand column), each coloured rectangle is a specific *MSP2* genotype. When the same genotype is detected over multiple timepoints, the same colour and name are used. For example, in the persistent infection number 1, the same genotype was detected in Dec, Jan, Feb, Mar and Apr (no information for May). In some volunteers, novel genotypes appear during the dry season, potentially indicating a new infection. However, in every persistent infection, at least one specific *MSP2* genotype is detected repeatedly throughout the dry season, showing that *P. falciparum* may remain asymptomatic for at least 6 months. Among persistent infections, a change in number of detected genotypes from one timepoint to the next did not associate with a change in parasitaemia, indicating that we probably detected most/all genotypes present in the infection.

## Supplementary Tables

**Table S1:** Details of antigens used in Luminex assay.

| Gene ID | Acronym | Description | Location | Expression tag | Strain | Reference |
| --- | --- | --- | --- | --- | --- | --- |
| PF3D7_0304600 | CSP | Most predominant and antigenic protein on sporozoite surface. Component of RTS,S vaccine | Sporozoite | n/a | 3D7 | Kastenmuller et al (PMID: 23275094) |
| PF3D7_1301600 | EBA140 RIII-V | erythrocyte binding antigen 140; involved in erythrocyte invasion | Apical organelles, micronemes | GST | 3D7 | Richards et al (PMID: 20843207) |
| PF3D7_0731500 | EBA175RII_F2 | erythrocyte binding antigen 175; RBC binding region via glycophorin A | Apical tip | GST | 3D7 | Richards et al (PMID: 20843207) |
| PF3D7_0102500 | EBA181 RIII-V | erythrocyte binding antigen 181; involved in erythrocyte invasion | Apical tip | GST | 3D7 | Richards et al (PMID: 20843207) |
| PF3D7_1035300 | GLURP R2 | Glutamate rich protein R2 | Merozoite Surface | n/a | F32 | Theisen et al (PMID: 7719909) |
| PF3D7_0206800 | MSP2 CH150/9 | CH150/9 allele of MSP2. Full-length. | Merozoite surface | GST | CH150/9 | Polley et al (PMID: 12654798) |
| PF3D7_0206800 | MSP2 Dd2 | Dd2 allele of MSP2. Full-length. | Merozoite surface | GST | DD2 | Taylor et al (PMID: 7591074) |
| PF3D7_1133400 | AMA1 | Apical membrane antigen 1 | Micronemes | His | FVO | Collins et al (PMID: 17192270) |
| PF3D7_0930300 | MSP1-19 | 19kDa fragment of MSP1 molecule. | Merozoite surface | GST | Wellcome | Burghaus and Holder (PMID: 8078519) |
| PF3D7_1335400 | Rh2_2030 | Reticulocyte-binding protein homolog 2; involved in erythrocyte invasion | Merozoites; Rhoptries | GST | 3D7 | Triglia et al (PMID: 11160005); Tetteh K unpublished |
| PF3D7_0424200 | Rh4.2 | Reticulocyte-binding protein homolog 4; involved in erythrocyte invasion | Merozoites; Rhoptries | GST | 3D7 | Reiling 2012 (PMID: 23028883); Tetteh K unpublished |
| PF3D7_0424100 | Rh5.1 | Reticulocyte-binding protein homolog 5; involved in erythrocyte invasion | Merozoites; Rhoptries | His | 3D7 | Hjerrild et al (PMID: 27457156); Tetteh K unpublished |
| PF3D7_0423700 | Etramp 4 Ag 2 | Early transcribed membrane antigen. Integral PVM protein. C-terminal | iRBC, PVM | GST | 3D7 | Helb et al (PMID: 26216993); Tetteh unpublished |
| PF3D7_0532100 | Etramp 5 Ag 1 | Early transcribed membrane antigen. Integral PVM protein | iRBC, PVM | GST | 3D7 | Spielmann et al (PMID: 12686607); Tetteh K unpublished |
| PF3D7_0501100.1 | HSP40 Ag 1 | Heat shock protein 40 | iRBC | GST | 3D7 | Helb et al (PMID: 26216993); Tetteh unpublished |
| PF3D7_1021800 | PfSEA-1 | Schizont egress antigen | iRBC | GST | 3D7 | Raj et al (PMID: 24855263); Tetteh K unpublished |
| PF3D7_0501300 | SBP1 | Skeleton-binding protein; translocation of PfEMP1 to RBC surface via Maurer's cleft | iRBC | GST | 3D7 | Gruring et al (PMID: 21266965); Tetteh K unpublished |

**Table S2:** Group characteristics.

|  | Short-lived | Persistent | Treated |
| --- | --- | --- | --- |
| **Haemoglobin type** |  |  |  |
| HbAA | 75% (12/16) | 79% (15/19) | 50% (1/2) |
| HbAS | 19% (3/16) | 21% (4/19) | 50% (1/2) |
| HbSS | 6% (1/16) |  |  |
| Concentration (mean g/dL) | 11.35 | 11.18 | 11.3 |
| **Age** |  |  |  |
| <5 | 3/17(18%) | 0/22 (0%) | 1/3 (33%) |
| 5-15 | 5/17 (29%) | 12/22 (55%) | 2/3 (67%) |
| >15 | 9/17 (53%) | 10/22 (45%) | 0/3 (0%) |
| **Sex** |  |  |  |
| Male | 8/17 (47%) | 12/22 (55%) | 2/3 67%) |
| Female | 9/17 (53%) | 10/22 (45%) | 1/3 (33%) |
| **Other** |  |  |  |
| Bednet usage (yes/total) | 36% (5/14) | 53% (10/19) | 66% (2/3) |
| Indoor Residual Spraying (yes/total) | 7% (1/14) | 0% (0/19) | 33% (1/3) |

## Funding

This work was supported by grants from the Netherlands Organization for Scientific Research (Vidi fellowship NWO 016.158.306) and the Bill & Melinda Gates Foundation (INDIE OPP1173572), the joint MRC/LSHTM fellowship, the French National Research Agency (18-CE15-0009-01).

## Conflict of interest

## Acknowledgments

We would like to thank Julia Mwesigwa and all fieldworkers for sample collection.

## Notes

### Competing Interest Statement

The authors have declared no competing interest.

### Author Declarations

The study was approved by the Gambia Government/Medical Research Council Joint Ethics Committee (SCC 1318, L2014.67, SCC 1472).

## References

1. Babiker HA: Unstable malaria in Sudan: the influence of the dry season. Plasmodium falciparum population in the unstable malaria area of eastern Sudan is stable and genetically complex. Trans R Soc Trop Med Hyg 1998, 92:585–589.

2. Ouedraogo AL, Goncalves BP, Gneme A, Wenger EA, Guelbeogo MW, Ouedraogo A, Gerardin J, Bever CA, Lyons H, Pitroipa X, et al: Dynamics of the Human Infectious Reservoir for Malaria Determined by Mosquito Feeding Assays and Ultrasensitive Malaria Diagnosis in Burkina Faso. J Infect Dis 2016, 213:90–99.

3. Mwesigwa J, Okebe J, Affara M, Di Tanna GL, Nwakanma D, Janha O, Opondo K, Grietens KP, Achan J, D’Alessandro U: On-going malaria transmission in The Gambia despite high coverage of control interventions: a nationwide cross-sectional survey. Malar J 2015, 14:314.

4. Bousema T, Okell L, Felger I, Drakeley C: Asymptomatic malaria infections: detectability, transmissibility and public health relevance. Nat Rev Microbiol 2014, 12:833–840.

5. Ashley EA, White NJ: The duration of Plasmodium falciparum infections. Malar J 2014, 13:500.

6. Hamad AA, El Hassan IM, El Khalifa AA, Ahmed GI, Abdelrahim SA, Theander TG, Arnot DE: Chronic Plasmodium falciparum infections in an area of low intensity malaria transmission in the Sudan. Parasitology 2000, 120 (Pt 5):447–456.

7. Andolina C, Rek JC, Briggs J, Okoth J, Musiime A, Ramjith J, Teyssier N, Conrad M, Nankabirwa JI, Lanke K, et al: Sources of persistent malaria transmission in a setting with effective malaria control in eastern Uganda: a longitudinal, observational cohort study. Lancet Infect Dis 2021.

8. Gupta S, Snow RW, Donnelly CA, Marsh K, Newbold C: Immunity to non-cerebral severe malaria is acquired after one or two infections. Nat Med 1999, 5:340–343.

9. Snow RW, Omumbo JA, Lowe B, Molyneux CS, Obiero JO, Palmer A, Weber MW, Pinder M, Nahlen B, Obonyo C, et al: Relation between severe malaria morbidity in children and level of Plasmodium falciparum transmission in Africa. Lancet 1997, 349:1650–1654.

10. Baird JK: Host age as a determinant of naturally acquired immunity to Plasmodium falciparum. Parasitol Today 1995, 11:105–111.

11. Andrade CM, Fleckenstein H, Thomson-Luque R, Doumbo S, Lima NF, Anderson C, Hibbert J, Hopp CS, Tran TM, Li S, et al: Increased circulation time of Plasmodium falciparum underlies persistent asymptomatic infection in the dry season. Nat Med 2020, 26:1929–1940.

12. Portugal S, Tran TM, Ongoiba A, Bathily A, Li S, Doumbo S, Skinner J, Doumtabe D, Kone Y, Sangala J, et al: Treatment of Chronic Asymptomatic Plasmodium falciparum Infection Does Not Increase the Risk of Clinical Malaria Upon Reinfection. Clin Infect Dis 2017, 64:645–653.

13. Crompton PD, Kayala MA, Traore B, Kayentao K, Ongoiba A, Weiss GE, Molina DM, Burk CR, Waisberg M, Jasinskas A, et al: A prospective analysis of the Ab response to Plasmodium falciparum before and after a malaria season by protein microarray. Proc Natl Acad Sci U S A 2010, 107:6958–6963.

14. Briggs J, Teyssier N, Nankabirwa JI, Rek J, Jagannathan P, Arinaitwe E, Bousema T, Drakeley C, Murray M, Crawford E, et al: Sex-based differences in clearance of chronic Plasmodium falciparum infection. Elife 2020, 9.

15. Gonzales SJ, Reyes RA, Braddom AE, Batugedara G, Bol S, Bunnik EM: Naturally Acquired Humoral Immunity Against Plasmodium falciparum Malaria. Front Immunol 2020, 11:594653.

16. Langhorne J, Ndungu FM, Sponaas AM, Marsh K: Immunity to malaria: more questions than answers. Nat Immunol 2008, 9:725–732.

17. Rono J, Osier FH, Olsson D, Montgomery S, Mhoja L, Rooth I, Marsh K, Farnert A: Breadth of anti-merozoite antibody responses is associated with the genetic diversity of asymptomatic Plasmodium falciparum infections and protection against clinical malaria. Clin Infect Dis 2013, 57:1409–1416.

18. Wu L, Mwesigwa J, Affara M, Bah M, Correa S, Hall T, Singh SK, Beeson JG, Tetteh KKA, Kleinschmidt I, et al: Antibody responses to a suite of novel serological markers for malaria surveillance demonstrate strong correlation with clinical and parasitological infection across seasons and transmission settings in The Gambia. BMC Med 2020, 18:304.

19. Achan J, Reuling IJ, Yap XZ, Dabira E, Ahmad A, Cox M, Nwakanma D, Tetteh K, Wu L, Bastiaens GJH, et al: Serologic Markers of Previous Malaria Exposure and Functional Antibodies Inhibiting Parasite Growth Are Associated With Parasite Kinetics Following a Plasmodium falciparum Controlled Human Infection. Clin Infect Dis 2020, 70:2544–2552.

20. Taylor SM, Parobek CM, Fairhurst RM: Haemoglobinopathies and the clinical epidemiology of malaria: a systematic review and meta-analysis. Lancet Infect Dis 2012, 12:457–468.

21. White NJ: Anaemia and malaria. Malar J 2018, 17:371.

22. Akiyama T, Pongvongsa T, Phrommala S, Taniguchi T, Inamine Y, Takeuchi R, Watanabe T, Nishimoto F, Moji K, Kano S, et al: Asymptomatic malaria, growth status, and anaemia among children in Lao People’s Democratic Republic: a cross-sectional study. Malar J 2016, 15:499.

23. Hofmann N, Mwingira F, Shekalaghe S, Robinson LJ, Mueller I, Felger I: Ultra-sensitive detection of Plasmodium falciparum by amplification of multi-copy subtelomeric targets. PLoS Med 2015, 12:e1001788.

24. Meerstein-Kessel L, Andolina C, Carrio E, Mahamar A, Sawa P, Diawara H, van de Vegte-Bolmer M, Stone W, Collins KA, Schneider P, et al: A multiplex assay for the sensitive detection and quantification of male and female Plasmodium falciparum gametocytes. Malar J 2018, 17:441.

25. Mueller I, Schoepflin S, Smith TA, Benton KL, Bretscher MT, Lin E, Kiniboro B, Zimmerman PA, Speed TP, Siba P, Felger I: Force of infection is key to understanding the epidemiology of Plasmodium falciparum malaria in Papua New Guinean children. Proc Natl Acad Sci U S A 2012, 109:10030–10035.

26. Grignard L, Mair C, Curry J, Mahey L, Bastiaens GJH, Tiono AB, Okebe J, Coulibaly SA, Goncalves BP, Affara M, et al: Bead-based assays to simultaneously detect multiple human inherited blood disorders associated with malaria. Malar J 2019, 18:14.

27. Wu L, Hall T, Ssewanyana I, Oulton T, Patterson C, Vasileva H, Singh S, Affara M, Mwesigwa J, Correa S, et al: Optimisation and standardisation of a multiplex immunoassay of diverse Plasmodium falciparum antigens to assess changes in malaria transmission using sero-epidemiology. Wellcome Open Res 2019, 4:26.

28. Bejon P, Andrews L, Hunt-Cooke A, Sanderson F, Gilbert SC, Hill AV: Thick blood film examination for Plasmodium falciparum malaria has reduced sensitivity and underestimates parasite density. Malar J 2006, 5:104.

29. Ahmad A, Prom A, Bradley J, Ndiath M, Etoketim B, Bah M, Van Geertruyden JP, Drakeley C, Bousema T, Achan J, D’Alessandro U: Gametocyte carriage after seasonal malaria chemoprevention in Plasmodium falciparum infected asymptomatic children. Malar J 2021, 20:169.

30. Buchwald AG, Sorkin JD, Sixpence A, Chimenya M, Damson M, Wilson ML, Seydel K, Hochman S, Mathanga D, Taylor TE, Laufer MK: Association Between Age and Plasmodium falciparum Infection Dynamics. Am J Epidemiol 2019, 188:169–176.

31. Bretscher MT, Maire N, Felger I, Owusu-Agyei S, Smith T: Asymptomatic Plasmodium falciparum infections may not be shortened by acquired immunity. Malar J 2015, 14:294.

32. Okebe J, Affara M, Correa S, Muhammad AK, Nwakanma D, Drakeley C, D’Alessandro U: School-based countrywide seroprevalence survey reveals spatial heterogeneity in malaria transmission in the Gambia. PLoS One 2014, 9:e110926.

33. Nassir E, Abdel-Muhsin AM, Suliaman S, Kenyon F, Kheir A, Geha H, Ferguson HM, Walliker D, Babiker HA: Impact of genetic complexity on longevity and gametocytogenesis of Plasmodium falciparum during the dry and transmission-free season of eastern Sudan. Int J Parasitol 2005, 35:49–55.

34. Smith T, Beck HP, Kitua A, Mwankusye S, Felger I, Fraser-Hurt N, Irion A, Alonso P, Teuscher T, Tanner M: Age dependence of the multiplicity of Plasmodium falciparum infections and of other malariological indices in an area of high endemicity. Trans R Soc Trop Med Hyg 1999, 93 Suppl 1:15–20.

35. Nkhoma SC, Trevino SG, Gorena KM, Nair S, Khoswe S, Jett C, Garcia R, Daniel B, Dia A, Terlouw DJ, et al: Co-transmission of Related Malaria Parasite Lineages Shapes Within-Host Parasite Diversity. Cell Host Microbe 2020, 27:93–103 e104.

36. Koepfli C, Schoepflin S, Bretscher M, Lin E, Kiniboro B, Zimmerman PA, Siba P, Smith TA, Mueller I, Felger I: How much remains undetected? Probability of molecular detection of human Plasmodia in the field. PLoS One 2011, 6:e19010.

